# Low Transmission of the Globally Dominant Recombinant SARS-CoV-2 XFG variant in Kenya, May-July 2025

**DOI:** 10.1101/2025.11.10.25338449

**Authors:** Arnold W. Lambisia, Joyce Nyiro, Esther Nyadzua Katama, Doreen Lugano, Angela W. Maina, Martin Mutunga, Hillary Wafula, James Nyagwange, Philip Bejon, Simon Delicour, Edward C. Holmes, Isabella Ochola-Oyier, Charles J. Sande, Charles N. Agoti

## Abstract

**Objective:** The ongoing SARS-CoV-2 evolution has produced over 5,200 genetically distinct PANGO lineages whose epidemiological trajectories differ across regions. In early 2025, a new recombinant SARS-CoV-2 variant named XFG emerged, predominating in most global regions by June 2025. Here we describe XFG introduction and molecular epidemiological patterns in Kenya, May**-**July 2025.

**Results:** Of 7,564 nasopharyngeal/oropharyngeal swabs sampled across three surveillance platforms (a community cohort and two outpatient ARI) and tested by quantitative PCR, only 23 (0.03%) were positive. From these, we recovered six near-complete genomes that mapped to PANGO lineages XFG.12, XFG.4.1, XFG.7 and XFG.21. The median age of the positive cases was 18 years (IQR 10.0–21.8) and presented mainly with runny nose (47.6%), cough (42.9%), sore throat (33.3%) and fever (33.3%). Phylogenetic analysis including 14 sequences from Nairobi deposited on GISAID suggested at least 13 introduction events of the XFG variant into Kenya. Amino acid differences were observed between the Kenyan XFG.12 and XFG.4.1 in the ORF1a (H45Y) and ORF1b (D1848X/Y) proteins respectively. We confirm introduction but limited transmission of the XFG variant in Kenya during May–July 2025. This observation underlines the importance of regional genomic surveillance for appropriate and optimized intervention design.

## Background

On January 27, 2025, a new recombinant severe acute respiratory syndrome coronavirus 2 (SARS-CoV-2) variant named XFG, was first detected in Canada ^1^. Based on data available in Global Initiative on Sharing All Influenza Data (GISAID; October 4, 2025), the XFG variants appear to have spread globally (>88 countries; **Figure 1**) and were estimated to have a growth advantage of 27% as of October 2025 [defined as the percentage increase in a variant’s growth rate per week] compared to co-circulating variants, reflecting its intrinsic fitness ^2,3^.

**Figure 1.**
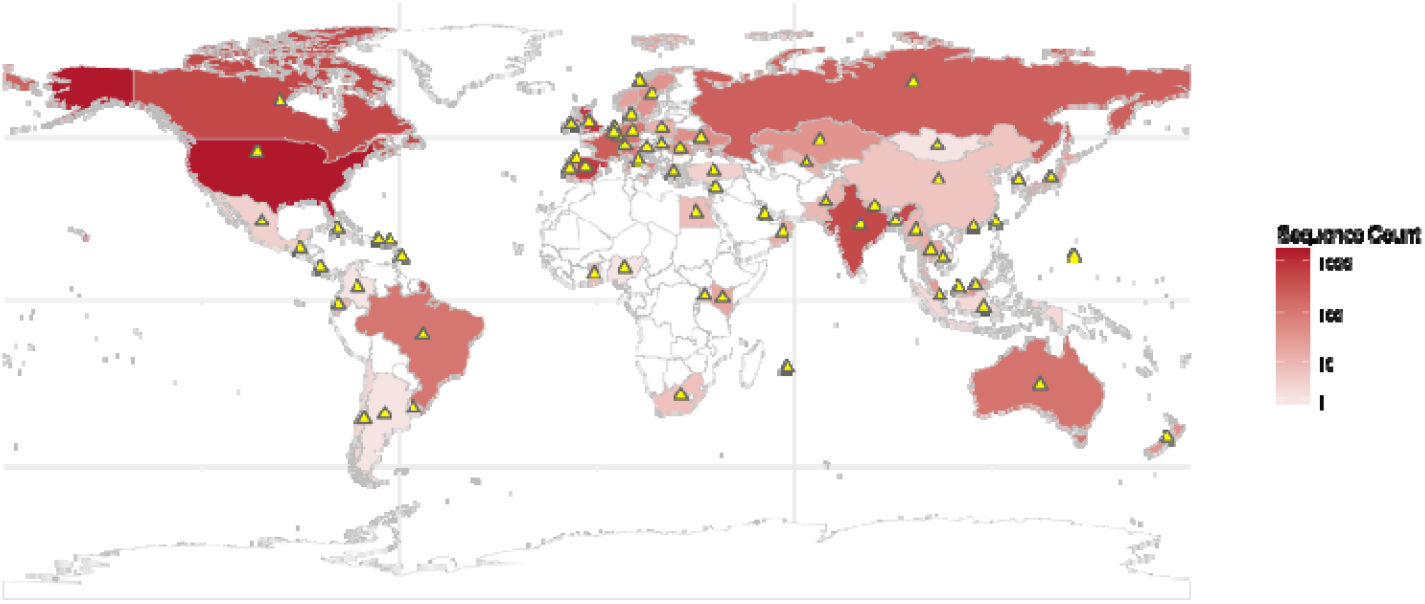
World map showing the spatial distribution of XFG SARS-CoV-2 sequences from GISAID. Color intensity corresponds to the number of sequences. The yellow triangles show countries that have detected XFG.12, XFG.21, XFG.4.1 and XFG.7 lineages also detected in Kenya

The XFG variant is a recombinant of the Omicron PANGO lineages LF.7 and LP.8.1.2 and has four key defining mutations in the spike (S) protein: His445Arg, Asn487Asp, Gln493Glu, and Thr572Ile. These changes have been demonstrated to enhance immune escape from class1/2 antibodies but are associated with lower angiotensin-converting enzyme 2 (ACE-2) binding capability and infectivity than PANGO lineage LP.8.1.1 ^3^. By September 22, 2025, 101 XFG PANGO lineages had been identified with 52 bearing lineage-defining spike (S) mutations either in the N-terminal domain (S1-NTD; n=18), receptor-binding domain (S1-RBD; n=11) or S2 (n=24) (https://mdu-phl.github.io/pango-watch/).

Like other locations globally, new SARS-CoV-2 waves of infection have continued to arise in Kenya, albeit with a lengthening inter-wave intervals and decreasing disease severity ^4^. To monitor its post emergency epidemiology, we have maintained SARS-CoV-2 surveillance in the various regions of Kenya through three diverse platforms i.e., pediatric inpatient surveillance, sentinel outpatient surveillance, and community-based homestead surveillance ^4,5^. Ten waves of infection have been recorded through these surveillance platforms to date ^4–7^. On the Kenyan coast, where most samples are taken, a wave was experienced between November 2024 and January 2025. Genomic sequencing showed this wave was predominated by the variant LF7.* ^4^. Following this period (Feb-July 2025) and coinciding with a period of heightened detections of the XFG variant globally, only sporadic cases of SARS-CoV-2 were detected from our surveillance. Here, we report molecular-epidemiological patterns of XFG variant detections in Kenya, May-July 2025.

## Methods

### Study design and participants

The design and populations of the underlying studies that provided the samples analyzed here have been previously described in detail elsewhere except for one (the Platform for Rapid Immunological Characterization Of SARS-CoV-2 variants in Kenya and the Eastern Africa Region i.e., PRICOS study) ^4,5^. Briefly, the analyzed nasopharyngeal/oropharyngeal (NP/OP) samples (collected in universal transport media; UTM) were part of; (a) Kilifi Health and Demographic Surveillance Systems (KHDSS) area Health Facility outpatient (KHDSS-HF study ^4^), (b) PRICOS study and (c) Respiratory Virus Reinfection study (ResViRe ^5^). Participants from the KHDSS-HF study presented with acute respiratory illness (ARI) to one of the five enrolled outpatient facilities within the KHDSS area ^4^. The PRICOS study participants presented with ARI in facilities within the counties of Bungoma, Nakuru, Naivasha, Nairobi and Machakos. The ReSViRe study is a community surveillance where ∼ 500 participants (residents of Kilifi County) are sampled at home 1-2 times a week regardless of the symptom status ^5^. Participants in all three studies were from across all age ranges.

### Laboratory procedures Diagnostics and sequencing

Total ribonucleic acids (RNA) were extracted from 140μl the NP/OP UTM using the automated RNeasy Mini Kit (QIAGEN, Manchester, UK). SARS-CoV-2 was screened in the RNA extracts the QIAGEN Multiplex RT-PCR + R Kit (QIAGEN, Manchester, UK) using primers/probe targeting the Envelope protein as previously described ^5^. Positive samples were considered as those with a cycle threshold (Ct) of <35.0 in experiments where both the positive and negative controls worked as expected. Whole genome sequencing was attempted for all positive samples using ARTIC primers v5.3.2 on the Oxford Nanopore Technologies (ONT) GridION instrument as previously described ^4,5^.

### Bioinformatic analysis

Consensus genomes were generated using the ARTIC bioinformatics pipeline (https://artic.readthedocs.io/en/latest/). Briefly, pod5 file were demultiplexed to generate fastq reads which were filtered based on length, mapped to a reference sequence (Genbank accession: MN908947.3), primer sequences trimmed, variants called and filtered, and a consensus genome generated with a minimum read depth of 20 reads. Genomic positions with lower than this minimum read depth were assigned ambiguous nucleotide (N). Assembled contigs with ≥70% genome coverage were assigned PANGO lineages^8^ using NextClade v.3.10.2 and the dataset “nextstrain/sars-cov-2/wuhan-hu-1” v.2025-09-09T12:13:13Z.

All global genomic data on XFG variant (n=24,706) was downloaded from GISAID on October 4, 2025, and used to compare the temporal trends of identified PANGO lineages in coastal Kenya and Nairobi Kenya with those globally. With the exception sequences collected in Kenya, the global dataset was subsampled to achieve ten sequences per country per month and per lineage for the locally identified lineages, generating a total data set of 1,789 genome sequences. A maximum likelihood (ML) and time-resolved tree were generated using IQTREE v2.1.3 assuming the general time reversible (GTR) model of nucleotide substitution and TreeTime v0.11.4 respectively. Import events into Kenya were estimated using TreeTime “mugration” model with the country of origin for the global data as discrete traits to reconstruct geographic locations at all internal nodes as previously described ^5^.

## Results

Between May 1, 2025, and July 31, 2025, we obtained 7,564 NP/OP swabs (ReSViRe study; n=6131, KHDSS-HF; n= 1013, PRICOS; n=420) from 1,935 individuals for SARS-CoV-2 screening. Of these 23 (0.03%) NP/OP swabs from 21 individuals were quantitative PCR positive for SARS-CoV-2 (5 from KHDSS-HF, 3 from PRICOS and 13 from ResViRe). These had a median Ct value of 33.0 (inter-quartile range (IQR) 28.6–34.6). The sampling platform distribution, demographic and clinical characteristics of the 18 positive individuals are summarized in **Table 1**. The median age of the positive cases was 13.5 years (IQR 8.7-20.5), and most common symptoms were a runny nose (38.9%), cough (33.3%), sore throat (33.3%) and fever (27.8%).

**Table 1.** Clinical and demographic characteristics of SARS-CoV-2 positive cases in Kilifi between May and July 2025.

|  | Outpatient Surveillance (n=5) | Community Surveillance (n=13) | Outpatient Surveillance non-KHDSS (n=3) | Total (n=21) |
| --- | --- | --- | --- | --- |
| **Sex** |  |  |  |  |
| Female | 2 (40.0%) | 8 (61.5%) | 1 (33.3%) | 11 (52.4%) |
| Male | 3 (60.0%) | 5 (38.5%) | 2 (66.7%) | 10 (47.6%) |
| **Age (years)** |  |  |  |  |
| Median (IQR) | 18.0 (10.0, 19.0) | 13.3 (8.3, 21.8) | 34.0 (27.0, 57.0) | 18.0 (10.0, 21.8) |
| **Age groups (years)** |  |  |  |  |
| 0-13 | 2 (40.0%) | 6 (46.2%) | 0 (0.0%) | 8 (38.1%) |
| 13-19 | 1 (20.0%) | 3 (23.1%) | 0 (0.0%) | 4 (19.0%) |
| >19 | 2 (40.0%) | 4 (30.8%) | 3 (100.0%) | 9 (42.9%) |
| **Cycle threshold value** |  |  |  |  |
| Median (IQR) | 32.4 (27.5, 34.5) | 33.5 (32.7, 34.7) | 28.2 (26.7, 30.4) | 33.0 (28.6, 34.6) |
| **Pango lineage** |  |  |  |  |
| XFG.12 | 2 (40.0%) | 0 (0.0%) | 0 (0.0%) | 2 (9.5%) |
| XFG.21 | 1 (20.0%) | 0 (0.0%) | 0 (0.0%) | 1 (4.8%) |
| XFG.4.1 | 1 (20.0%) | 1 (7.7%) | 0 (0.0%) | 2 (9.5%) |
| XFG.7 | 0 (0.0%) | 0 (0.0%) | 1 (33.3%) | 1 (4.8%) |
| Failed sequencing | 1 (20.0%) | 12 (92.3%) | 2 (66.7%) | 15 (71.4%) |
| **Symptom status** |  |  |  |  |
| asymptomatic | 0 (0.0%) | 11 (84.6%) | 0 (0.0%) | 11 (52.4%) |
| symptomatic | 5 (100.0%) | 2 (15.4%) | 3 (100.0%) | 10 (47.6%) |
| **Clinical characteristics** |  |  |  |  |
| Runny nose | 5 (100.0%) | 2 (15.4%) | 3 (100.0%) | 10 (47.6%) |
| Cough | 5 (100.0%) | 1 (7.7%) | 3 (100.0%) | 9 (42.9%) |
| Sore throat | 4 (80.0%) | 0 (0.0%) | 3 (100.0%) | 7 (33.3%) |
| Fever | 5 (100.0%) | 0 (0.0%) | 2 (66.7%) | 7 (33.3%) |
| Headache | 1 (20.0%) | 0 (0.0%) | 3 (100.0%) | 4 (19.0%) |
| Chest pain | 0 (0.0%) | 0 (0.0%) | 2 (66.7%) | 2 (9.5%) |

Near complete genomes (median length 97.1%; IQR 94.9%-97.7%) were recovered from six samples and these classified into XFG.12 (n=2), XFG.4.1 (n=2), XFG.7 (n=1) and XFG.21 (n=1). The failed sequencing samples had a low viral load (median Ct 33.5 (range: 32.5-34.8) *vs*. 30.3 (range: 27.7-32.5)). Five sequenced samples were from sentinel outpatient surveillance, and the remaining one was from community surveillance. The temporal lineage patterns of sequenced SARS-CoV-2 samples from Kilifi and Machakos between May 2025 and July 2025 is shown in **Figure 2A**. An additional 14 Kenyan sequences of samples from Nairobi, collected over the same period were available. These mapped to lineages XFG.12 (n=6) and XFG.3 (n=8) (**Figure 2B**).

**Figure 2.**
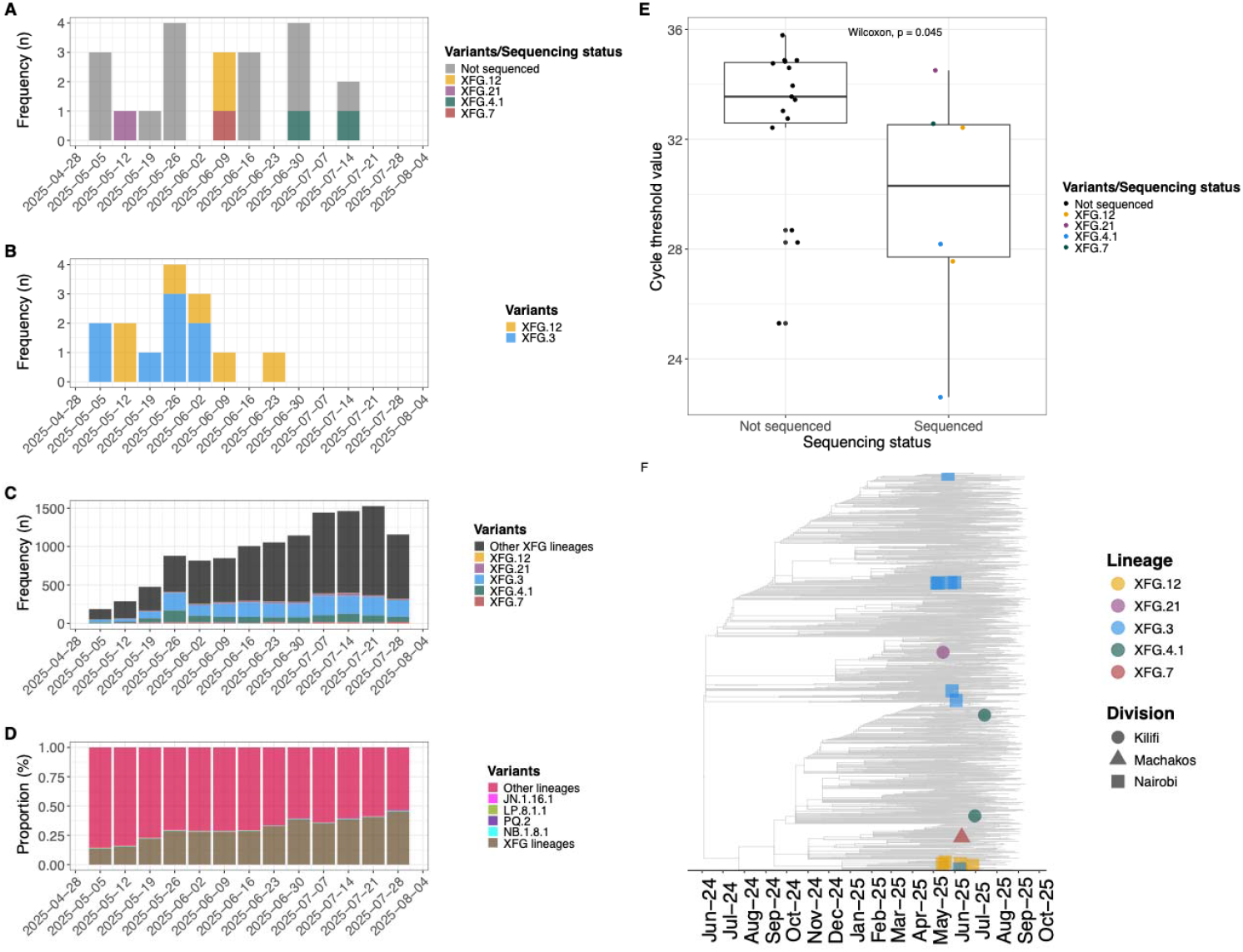
Weekly Local and global molecular epidemiological patterns of SARS-CoV-2 XFG variants between May and July 2025. **A)**. Weekly temporal distribution of the XFG variants detected in samples **(A)** from this study, **(B)** from Nairobi Kenya, and **(C)** global sequence data on GISAID. The bars are colored by the SARS-CoV-2 lineages. **(D)** Weekly distribution of 41,747 sequences from GISAID grouped as either XFG variant, other lineages, or lineages that surpassed the 5% weekly threshold over the study period. (**E**) A time-resolved phylogeny showing the clustering of Kenyan sequences collected during the surveillance period. The different tip colors indicate the different sub-variants detected and are shaped by the county of sample collection.

Among the Coastal Kenya observed PANGO lineages, XFG.3 and XFG.4.1 were commonly detected globally compared to XFG.12 and XFG.21 **(Figure 2C)**. At the end of July 2025, XFG lineages were ∼50% of the weekly collected and deposited sequences on GISAID (**Figure 2D**). Other PANGO lineages including JN.1.16.1, LP.8.1.1, PQ.2 and NB.1.8.1 surpassed a 5% threshold of weekly sequences deposited in GISAID between May and July 2025 (**Figure 2D**).

The phylogenetic relatedness of the Kenyan (Kilifi and Nairobi) and 1,790 global XFG* genomes is shown in **Figure 2F**. The Kenyan XFG sequences were distributed across multiple clusters interspersed by global sequences pointing towards multiple introductions of the variant into the country. Using the TreeTime “mugration” model we estimated thirteen introduction events of the XFG* variants in Kenya from USA, Canada, Spain, France, New Zealand and Denmark. Limited current genomic surveillance makes it a bit difficult to do finescale phylogeographic analysis as shown by the global distribution of all XFG data downloaded from GISAID in **Figure1**. Non-synonymous SNPs identified in Kenyan SARS-CoV-2 genomes included mutations in the XFG.12 lineage (ORF1a:H45Y, ORF1b:D1848Y, ORF1b:T1453I, M:F100L and E:N64K) and XFG.3 lineage (M:F100X, E:N64X and ORF7B:L32F/X).

## Discussion

The XFG variant was linked to the increase in SARS-CoV-2 positive cases globally in 2025 and was designated a variant under monitoring by WHO on June 25, 2025 ^1^.

We report its limited transmission in Kenya, May-July 2025. Despite screening 7,564 samples at the height of the XFG global epidemic, only 23 (0.03%) were found SARS-CoV-2 positive. Sequencing identified four XFG PANGO lineages each characterized by distinct amino acid substitutions: XFG.21 (ORF1b:S997L), XFG.12 (ORF1b:T1453I), XFG.4.1 (ORF1b:V2223L) and XFG.7(N:K65I, ORF9b:N62Y).

These limited numbers of XFG variant detections in coastal Kenya could be due to: (a) the high local population immunity against the variant potentially conferred by the closely related LF.7 variants that dominated wave 10 ^9^ (noting that LF.7 is one of the two lineages that recombined to give rise to XFG); and/or (b) the different XFG lineages detected in Kenya being distinguished by mutations in the ORF1b region rather than the S protein. This may explain their limited sustained transmission as mutations in the ORF1 do not lead to drastic transmission changes. Ongoing surveillance will inform if the XFG detections will increase. in subsequent months.

Notably, our analysis also reveals that the XFG recombinant variant was introduced into Kenya multiple times. Similar findings of repeated variant introductions has appeared across all previous waves of along the Kenya coast ^5,6^. These findings support the notion that new SARS-CoV-2 variants are continually introduced into the local population, with outbreaks contingent on local factors such as population immunity, co-circulating respiratory viruses (virus–virus interactions), climatic seasons, and social determinants such as school term status, national/religious events etc.

As well as XFG, multiple globally dominant SARS-CoV-2 lineages have had limited establishment in coastal Kenya (e.g., XBB.1.5, EG.5.1.1, HV.1, JN.1, KP.3.1 and XEC). This may be due to the region’s unique SARS-CoV-2 exposure history caused by unique locally dominant lineages (e.g., FY.4, GE.1.2, XBB.2.3, and LP.7). ^4,5^. The frequent mismatch between locally dominant and globally circulating strains implies that the protection efficiency of vaccines selected based on globally dominant strains may not be optimal locally. Our study emphasizes regional genomic surveillance is critical to inform appropriate local interventions.

This study had limitations. First, participants were drawn from a few regions in Kenya. As a result, sampling bias may exist if the variant circulated more extensively outside these areas. Second, we have not assessed the immune recognition of this variant in the local population, so we cannot confirm inability to escape local immunity. Third, sequencing success was limited, likely due to low viral loads (high Ct values, **Figure 2E**). Finally, we cannot make inferences about the clinical spectrum/severity of XFG infections in this population due to small patient numbers. Nevertheless, we think these limitations are unlikely to alter our study conclusions.

In conclusion, in the post-emergency period, ongoing *SARS-CoV-2*genomic surveillance has continued to reveal the emergence of novel variants that can rapidly spread globally. XFG, a recombinant variant, reached global dominance by mid-2025. Its period of high activity coincided with the low season in Kenya. Although evidence suggests multiple introductions of XFG into Kenya, these events have not yet resulted in large outbreaks. Looking ahead, integrated studies combining genomic, immunological, virological, and epidemiological data will be essential to illuminate the factors that determine which variants dominate locally and globally, for more effective outbreak control.

## Abbreviations

CI: Confidence interval
Ct: Cycle threshold
IQR: interquartile range
GISAID: Global Initiative on Sharing All Influenza Data
HF: Health facility
KHDSS: Kilifi Health and Demographic Surveillance
ML: Maximum likelihood
NP/OP: Nasalpharyngeal/oropharyngeal
ORF: Open reading frame
UTM: Universal Transport Media
PANGO: Phylogenetic Assignment of Named Global outbreak Lineages
qPCR: Quantitative polymerase chain reaction
SARS-CoV-2: Severe acute respiratory syndrome coronavirus 2

## Declarations

### Ethics approval and consent to participate

The protocols’ consenting and sample collection process were reviewed and approved by KEMRI Scientific Ethics and Research Unit (SERU), Nairobi Kenya (protocol numbers #3103, #4724 and #4807).

### Consent for publication

Not applicable.

### Availability of data and materials

Consensus SARS-CoV-2 genomes of the sequenced samples in this study have been deposited in the Global Initiative on Sharing all Influenza Data (GISAID) database and can be accessed at https://doi.org/10.55876/gis8.250922yh.

Patient demographic and clinical data and scripts that were used for data analysis in this study are accessible on the Harvard Dataverse using the link https://doi.org/10.7910/DVN/LJEYL6.

## Author contributions

A.W.L, D.L, A.W.M, M.M and H.W were involved in investigation. C.N.A acquired the funds for the study and conceptualized the study. C.N.A, J.N and C.S were involved in supervision. A.W.L and C.N.A wrote the original draft, reviewed and edited the manuscript. P.B, G.G, S.D, L.I.O, E.C.H were involved in writing – reviewing and editing. A.W.L and E.K were involved in data curation and visualization. All authors reviewed the manuscript.

## Acknowledgments

We thank members of the Pathogen Epidemiology and Omics (PEO) Group particularly the field team who collected the clinical samples and metadata and laboratory diagnostics team who processed the samples analysed and presented in this report. We thank all genomic data contributors including authors and their originating labs of the sequence data in GISAID that we included in this research (https://doi.org/10.55876/gis8.250922om). This working is published with permission from director KEMRI.

## References

1. WHO. WHO TAG-VE Risk Evaluation for SARS-CoV-2 Variant Under Monitoring: XFG Executive Summary [Internet]. 2025 Jun [cited 2025 Sep 17]. Available from: https://www.who.int/docs/default-source/coronaviruse/25062025_xfg_ire.pdf

2. Chen C, Nadeau S, Yared M, Voinov P, Xie N, Roemer C, et al. CoV-Spectrum: analysis of globally shared SARS-CoV-2 data to identify and characterize new variants. Bioinformatics. 2022 Mar 4;38(6):1735–7.

3. Guo C, Yu Y, Liu J, Jian F, Yang S, Song W, et al. Antigenic and virological characteristics of SARS-CoV-2 variants BA.3.2, XFG, and NB.1.8.1. Lancet Infect Dis. 2025 Jul;25(7):e374–7.

4. Lambisia AW, Nyiro J, Githinji G, Katama EN, Moraa E, Mwita JM, et al. Longitudinal Epidemiology and Variant Dynamics of SARS-CoV-2 in Coaystal Kenya (20202025): Clinical Features and Wave Patterns. medRxiv [Internet]. 2025; Available from: https://www.medrxiv.org/content/early/2025/08/26/2025.08.22.25334233

5. Lambisia AW, Katama EN, Moraa E, Mwita JM, Gallagher K, Mutunga M, et al. Genomic and clinical epidemiology of SARS-CoV-2 in coastal Kenya: insights into variant circulation, reinfection, and multiple lineage importations during a post-pandemic wave. BMC Global and Public Health [Internet]. 2025 Sep 9;3(1):80. Available from: https://bmcglobalpublichealth.biomedcentral.com/articles/10.1186/s44263-025-00201-6

6. Agoti CN, Ochola-Oyier LI, Dellicour S, Mohammed KS, Lambisia AW, de Laurent ZR, et al. Transmission networks of SARS-CoV-2 in Coastal Kenya during the first two waves: A retrospective genomic study. Elife [Internet]. 2022 Jun 14;11 (June 2022). Available from: 10.7554/eLife.71703

7. Githinji G, Lambisia AW, Omah I, O’Toole A, Mohamed KS, de Laurent ZR, et al. The genomic epidemiology of SARS-CoV-2 variants of concern in Kenya. medRxiv [Internet]. 2022 Jan 1;2022.10.26.22281446. Available from: http://medrxiv.org/content/early/2022/10/27/2022.10.26.22281446.abstract

8. Lugano D, Kutima B, Kimani M, Sigilai A, Gitonga J, Karani A, et al. Evaluation of population immunity against SARS-CoV-2 variants, EG.5.1, FY.4, BA.2.86, JN.1, JN.1.4, and KP.3.1.1 using samples from two health demographic surveillance systems in Kenya. BMC Infect Dis [Internet]. 2024 Dec 28;24(1):1474. Available from: https://bmcinfectdis.biomedcentral.com/articles/10.1186/s12879-024-10367-3

